# Alcohol consumption and telomere length: observational and Mendelian randomization approaches

**DOI:** 10.1101/2021.09.17.21263720

**Authors:** Anya Topiwala, Bernd Taschler, Klaus P. Ebmeier, Steve Smith, Hang Zhou, Daniel F Levey, Veryan Codd, Nilesh Samani, Joel Gelernter, Thomas E. Nichols, Stephen Burgess

## Abstract

Alcohol’s impact on telomere length, a proposed marker of biological age, is unclear. We performed the largest observational study to date and compared findings with Mendelian randomization (MR) estimates. Two-sample MR used data from a recent genome-wide association study (GWAS) of telomere length. Genetic variants were selected on the basis of associations with alcohol consumption and alcohol use disorder (AUD). Non-linear MR employed UK Biobank individual data. MR analyses suggest a causal relationship between alcohol and telomere length: both genetically predicted alcohol traits were inversely associated with telomere length. 1 S.D. higher genetically-predicted log-transformed alcoholic drinks weekly had a -0.07 S.D. effect on telomere length (95% confidence interval [CI]:-0.14 to -0.01); genetically-predicted AUD - 0.06 S.D. effect (CI:-0.10 to -0.02). Results were consistent across methods and independent from smoking. Non-linear analyses indicated a potential threshold relationship between alcohol and telomere length. Our findings have implications for potential aging-related disease prevention strategies.

## Introduction

Telomere length is considered a potential biological marker of aging [1]. These repetitive nucleotide sequences, together with associated protein complexes, form a ‘cap’ at the ends of chromosomes, protecting them from damage. As a cell’s replicative machinery cannot completely copy the ends of chromosomes, 50-100 base pairs are lost at each division. Telomere attrition therefore occurs with increasing cellular age. Critically short telomeres trigger cell death or replicative senescence, or occasionally continued division, mutation and genetic aberrations. Epidemiologically, shorter leucocyte telomere length (LTL) has been linked to several aging-related diseases including Alzheimer’s disease, cancer and coronary artery disease [2-4]. Telomere length is partly heritable [4] and linked to sex [5], ethnicity and paternal age [6], but has also been linked to environmental and lifestyle factors, including exercise [7], smoking [8] and alcohol consumption.

Observational studies of the relationship of alcohol use to telomere length have produced conflicting results. The largest such study to date, of 4567 individuals, found no association between alcohol intake and either baseline or longitudinal change in telomere length [9]. Another analysis of two American cohorts (n=2623) also reported null findings [10]. On the other hand, a few small studies (sample size range, 255-1800) have observed associations with heavy drinking or alcohol use disorder (AUD). Participants with AUD have been reported to have shorter telomeres compared to healthy controls [11]. A longitudinal study of Helsinki businessmen observed higher midlife alcohol consumption to be associated with shorter telomere length in older age [12]. Drinking >30g/day of alcohol in older participants was associated with shorter telomeres in a Korean study [13]. Associations were stronger in those experiencing the alcohol flush reaction, raising the intriguing possibility that acetaldehyde, ethanol’s toxic breakdown product, is mechanistically involved. In a recent review of 27 studies, ten showed significant associations between alcohol use and telomere length [14]. However heterogeneity between studies in quantification of telomere length and categorization of alcohol intake hindered meta-analysis and aggregation of the data.

Mendelian randomization (MR) seeks to identify potentially causal determinants of an outcome. It estimates the association between genetically predicted levels of an exposure and an outcome of interest. Residual confounding and reverse causation aim to be less of a concern than in most other methods of analysis of data from observational studies [15]. Using MR, genetic proxies can be used to study the effects of genetically predicted variability in alcohol consumption or AUD risk. To our knowledge, no MR study of alcohol and telomere length has yet been attempted.

We conducted a large observational study of two alcohol phenotypes, alcohol consumption and AUD, and leucocyte telomere length (LTL). Linear and non-linear models were fitted. We then performed linear MR analyses to investigate the evidence for a causal effect between alcohol consumption/AUD and LTL. Estimates generated by our observational and genetic methods were compared.

Genetic distinction between different alcohol use traits motivates their separate analysis. Quantity/frequency measures such as drinks per week and AUDIT-C, while moderately genetically correlated with AUD, have distinct patterns of genetic correlation with other traits [16]. Furthermore, as there has been much speculation about potential J-shaped relationships between alcohol and health outcomes [17], we performed a non-linear MR analysis to examine the shape of the relationship between alcohol consumption and telomere length. Multiple robust methods were employed to test MR assumptions. These included use of non-drinkers as negative controls, testing one of the key assumptions that genetic proxies only impact an outcome via the exposure. Given the widespread exposure to alcohol across the world, clarification of any potential causal impact on telomere length is important.

## Methods

All analyses were performed in R (version 3.6.0) unless otherwise specified.

### Study population

Participants were drawn from UK Biobank [18], a prospective cohort study which recruited ∼500,000 volunteers aged 40-69 years in 2006-10. Data used in this study included self-reported alcohol consumption, biological samples (blood) for genetic analysis, and long-term follow-up through hospital record linkage. UKB genetic data (SNPs) were generated from the Affymetrix Axiom UK Biobank array (∼450,000 individuals) and UK BiLEVE array (∼50,000 individuals) following extensive quality control [19]. Ancestry principal components were generated using loadings from high-confidence SNPs in the 1000 Genomes Cohort. SNP dosages for instrumental variants were extracted from UKB v3 imputed genotype data using qctool (version 2.0.7). Participants with solely European ancestry (defined by self-report and ancestral PCs) were included to avoid population stratification.

### Alcohol traits

We used two different alcohol trait definitions, to correspond to quantity and AUD. 1) Alcohol consumption was self-reported at baseline. Participants were categorized into current, never and previous drinkers. For current drinkers total estimated UK units consumed weekly were calculated by summing across beverage types as previously described [20]. 2) Alcohol use disorder (AUD) cases were defined by the presence of a relevant ICD-9 or ICD-10 code in linked NHS Hospital Episode Statistics.

### Telomere length measurements

Leucocyte telomere length (LTL) measurements were ascertained on DNA collected at the baseline assessment using a well-validated qPCR assay. Measurements were reported as a ratio of the telomere repeat number to single-copy gene (T/S ratio), which were then log-transformed to approximate the normal distribution. Multiple quality checks to control and adjust for technical factors were undertaken as described elsewhere [21]. To aid comparison with other datasets, z-standardized LTL values were used.

### Genetic variants

Genetically predicted alcohol consumption was ascertained using an instrument composed of 93 variants associated with alcohol consumption (drinks per week) with genomewide significance (GWS) in the largest published GWAS comprising 941,280 individuals [22]. Summary statistics were used that did not include data from UK Biobank participants (n=226,223) to avoid sample overlap, which can bias estimates towards observational associations [23]. All SNPs were associated with alcohol at GWS (p<5×10^−8^) and not in linkage disequilibrium (defined as r^2^>0.1). For AUD, 24 conditionally independent genetic variants were chosen from the largest published GWAS, comprised of European or European American individuals within the Million Veterans Program (MVP) and the Psychiatric Genomics Consortium (PGC) [24]. AUD cases were defined using ICD 9/10 codes within the MVP (n=45,995) and DSM-IV alcohol dependence within the PGC (n=11,569). Again all SNPs were associated with AUD at GWS (p<5×10^−8^). Genetic associations with LTL were obtained from the largest GWAS to date of telomere length, in 472,174 UKB participants [4].

### Statistical analysis

#### Observational analysis

Participants with complete alcohol quantity, telomere, and covariate data were included. Separate multiple linear regression models were used to assess the relationship between LTL (dependent variable) and 1) alcohol consumption and 2) AUD. Factors previously described to associate with LTL were included as covariates in analysis: age at baseline, sex, and ethnicity [4]. Alcohol consumption was fitted with linear and non-linear models to examine the shape of any alcohol-telomere relationship. The latter comprised two approaches. First, alcohol consumption was categorized into quantiles, second restricted cubic splines (RCS – 5 knots) were applied to alcohol intake. Non-linearity was formally tested with an F-test for equality of coefficients. To test the hypothesis that acetaldehyde is mechanistically involved in damage to telomeres, an interaction term between alcohol intake and *ADH1B* genotype (rs1229984) was included to test the hypothesis that higher levels of acetaldehyde are associated with shorter telomeres.

#### Genetic analyses

MR was used to obtain estimates for the association between genetically predicted alcohol consumption/AUD and telomere length. Both linear and non-linear MR analyses were undertaken. Linear MR can be performed using summary statistics and so we were able to harness the power from large consortia GWAS in two-sample designs. Non-linear MR necessitates individual (participant-level) data, and thus was undertaken within the UKB.

Two-sample MR analyses were conducted using MendelianRandomization package (version 0.5.1) and TwoSampleMR (version 0.5.6). Variants were harmonized between datasets, ensuring the association between SNPs and exposure and that between SNPs and the outcome reflected the same allele. Several MR methods were performed as broadly consistent results across methods strengthen the causal inference. Inverse variance weighted (IVW) analysis regresses the effect sizes of variant-telomere associations against effect sizes of the variant-alcohol associations. A random effects model was implemented. Scatter plots and leave-one-out analysis were used to evaluate influential outliers. The MR-Egger method uses a weighted regression with an unconstrained intercept to remove the assumption (in IVW) that all genetic variants are valid IVs. A non-zero intercept term in MR-Egger can be used as evidence of directional pleiotropy. The median MR method is also more resistant to pleiotropy. It takes the medial IV estimate from all included variants, and therefore is robust when up to 50% genetic variants are invalid. MR-PRESSO attempts to reduce outlier bias by performing estimation of causal estimates after removal of outliers [25].

We performed additional sensitivity analyses to assess robustness of the findings. Checking for reverse causality in the alcohol/AUD-telomere relationship was done using the MR Steiger directionality test [26], as well as repeating MR analyses inverting the exposure and outcomes. 85 SNPs (excluding n=5 palindromic SNPs) that are genome-wide significant for telomere length [4], had available associations with alcohol consumption, and 67 SNPs (excluding n=1 palindromic SNP) that had available associations with alcohol use disorder were used as instrumental variables to assess for a causal effect on alcohol. Differing availability of SNP associations reflects array differences between studies. We also conducted two-sample multivariable MR to test whether the causal effect of alcohol on telomere length was confounded or mediated by smoking. Genetic variant associations with smoking (cigarettes per day) were obtained from a large GWAS for the 93 SNPs used as instrumental variants for alcohol [22] and 18 SNPs used as instrumental variants for alcohol use disorder (n=6 had no available association statistics). Both IVW and MR Egger were conducted using MVMR in the MendelianRandomization package.

Nonlinear MR was undertaken by stratifying UKB subjects with complete alcohol, telomere and genetic data into 5 quantiles based on residual alcohol consumption. This was defined as alcohol consumption minus the genetic contribution to alcohol intake (IV-free exposure). A genetic risk score for alcohol consumption was created by weighting each SNP dosage by effect size (beta coefficient) obtained from the primary GWAS, and then summing across all SNPs. Stratifying directly on alcohol consumption could induce an association between the IV and outcome where there was none, invalidating MR assumptions. In each stratum a linear MR estimate of alcohol-telomere length was calculated using the ratio of coefficients method. To examine for the presence of a trend in the estimates a meta-regression of the stratum-specific estimates on the median value of alcohol in each stratum was performed.

To test the MR assumption that genetic variants only act upon telomere length through alcohol or AUD, we used never and previous drinkers in UKB as negative controls. MR analyses were performed separately in current, never and previous drinkers. If the assumption holds, there should be no association in non-drinkers. In all individual data MR analyses, age, sex and the top 10 ancestral principal components were included as covariates.

## Results

### Participant characteristics UK Biobank

298,628 UKB participants had complete data on alcohol intake, telomere length and covariates and were included in the observational analysis. The majority of participants were current drinkers, with only 5% (n=13,985) reported as never drinkers and 4% (n=12,104) previous drinkers. Never drinkers comprised a higher proportion of females, had fewer educational qualifications, lower smoking rates and lower levels of exercise than current drinkers (Table 1).

**Table 1:** Baseline characteristics of UK Biobank participants (n=298,628) included in observational analysis, according to alcohol status.

Participant characteristics UK Biobank
|  | Never drinkers<br>N=13,985 | Previous drinkers<br>N=12,104 | Current drinkers<br>N=272,481 |
| --- | --- | --- | --- |
| Age<sup>1</sup>, years | 56.8 (8.6) | 56.9 (8.0) | 56.3 (8.1) |
| Sex, females n(%) | 9771 (69.9) | 6425 (53.1) | 132007 (48.4) |
| TDI<sup>1,2</sup> | -0.4 (3.4) | -0.2 (3.5) | -1.6 (2.9) |
| Qualifications none | 3290 (23.5) | 2889 (23.9) | 35115 (12.9) |
| Degree<sup>3</sup> | 4112 (29.4) | 3470 (28.7) | 101351 (37.2) |
| Smoking never, n(%) | 11507 (82.3) | 5401 (44.6) | 143179 (52.5) |
| previous, n(%) | 1668 (11.9) | 4909 (40.6) | 101941 (37.4) |
| current, n(%) | 810 (5.8) | 1794 (14.8) | 27361 (10.0) |
| Systolic blood pressure<sup>1</sup>, mmHg | 138.6 (20.2) | 137.2 (19.7) | 139.9 (19.5) |
| ADH1B dose<sup>1,4</sup> | 1.9 (0.4) | 1.9 (0.3) | 1.9 (0.2) |
| Exercise,<sup>1</sup> MET minutes | 119.3 (104.1) | 123.3 (107.8) | 128.2 (100.7) |

<sup>1</sup> Mean (standard deviation)
<sup>2</sup>Townsend Deprivation Index – calculated on census data. Higher score indicates greater material deprivation.
<sup>3</sup>Selected educational qualification categories shown for brevity. Degree indicates any college or university degree.
<sup>4</sup> SNP dosage of C allele of rs129984 (MAF=0.03), associated with increased alcohol consumption compared to the T allele on a population level.

### Observational analysis

There was a significant observational association between high alcohol intake and shorter LTL (Figure 1). Compared to lowest quintile drinkers (<6 units (48g) weekly), highest quartile drinkers (>29 units weekly (232g)) had significantly shorter telomeres (beta= -0.05, 95% CI: -0.07 to -0.04, p=2×10^−16^). We found no evidence that this association was moderated by *ADH1B* genotype as indicated by non-significant interaction terms (p>0.08). Individuals with an ICD diagnosis of alcohol dependence (n=1632) in their linked clinical records had significantly shorter LTL compared with controls (beta= -0.15, 95% CI: -0.19 to -0.14, p=10×10^−15^).

**Figure 1:**
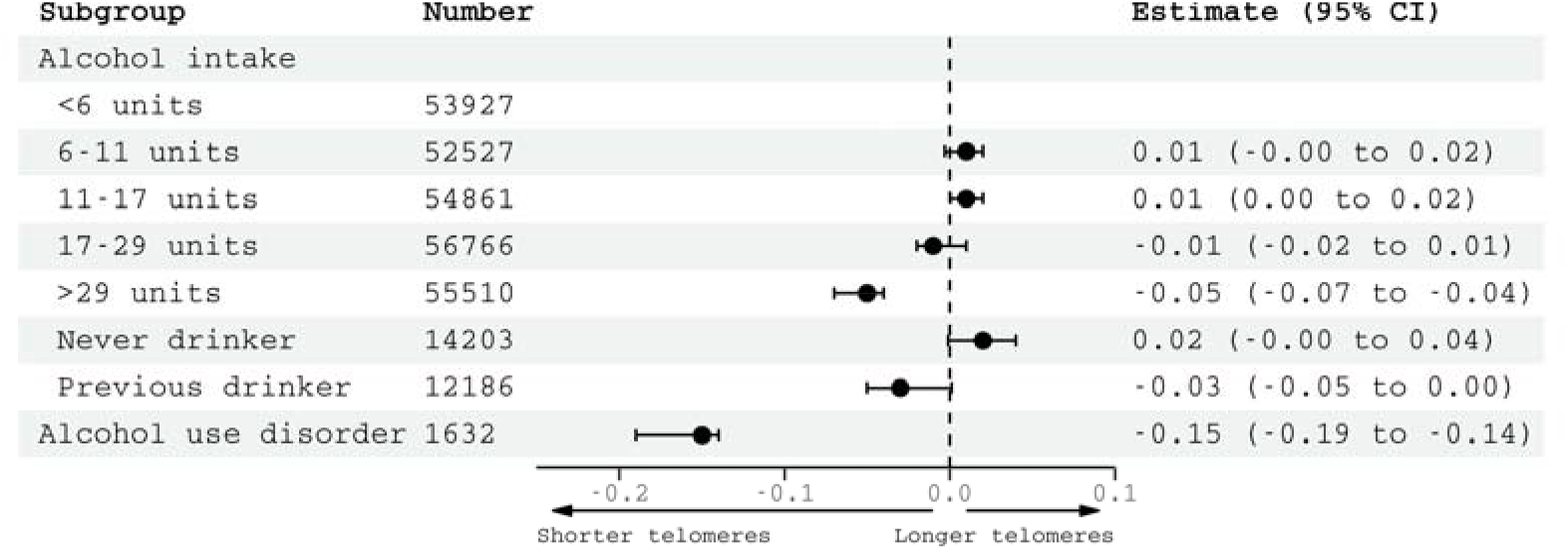
Observational associations with leucocyte telomere length in n=298,628 UK Biobank participants. Estimates generated from two regression models: 1) alcohol intake (estimates represent SD change in LTL for 1 S.D. increase in alcohol consumption) and 2) ICD diagnosis of AUD, plotted together for comparison. Reference category for alcohol intake is <6 units weekly. Models adjusted for: age, sex, ethnicity.

### Genetic analysis

Using 93 SNPs robustly and independently associated with alcohol consumption, univariable linear MR found an association between genetically predicted alcohol consumption and telomere length (IVW beta= -0.07, 95% CI: -0.14 to - 0.01, p=0.03) (Figure 2). Alternative methods gave consistent estimates, although there was some heterogeneity in estimates from different genetic variants (Figure 2). The strongest association was with the SNP rs1229984, in *ADH1B* an alcohol metabolism gene (supplementary Figure 5). This SNP explained the most variance in alcohol consumption (and AUD) of all variants. Excluding rs1229984 from the analyses slightly attenuated the MR IVW estimate which became insignificant (beta= -0.06, 95% CI= -0.13 to 0.01, p=0.09).

**Figure 2:**
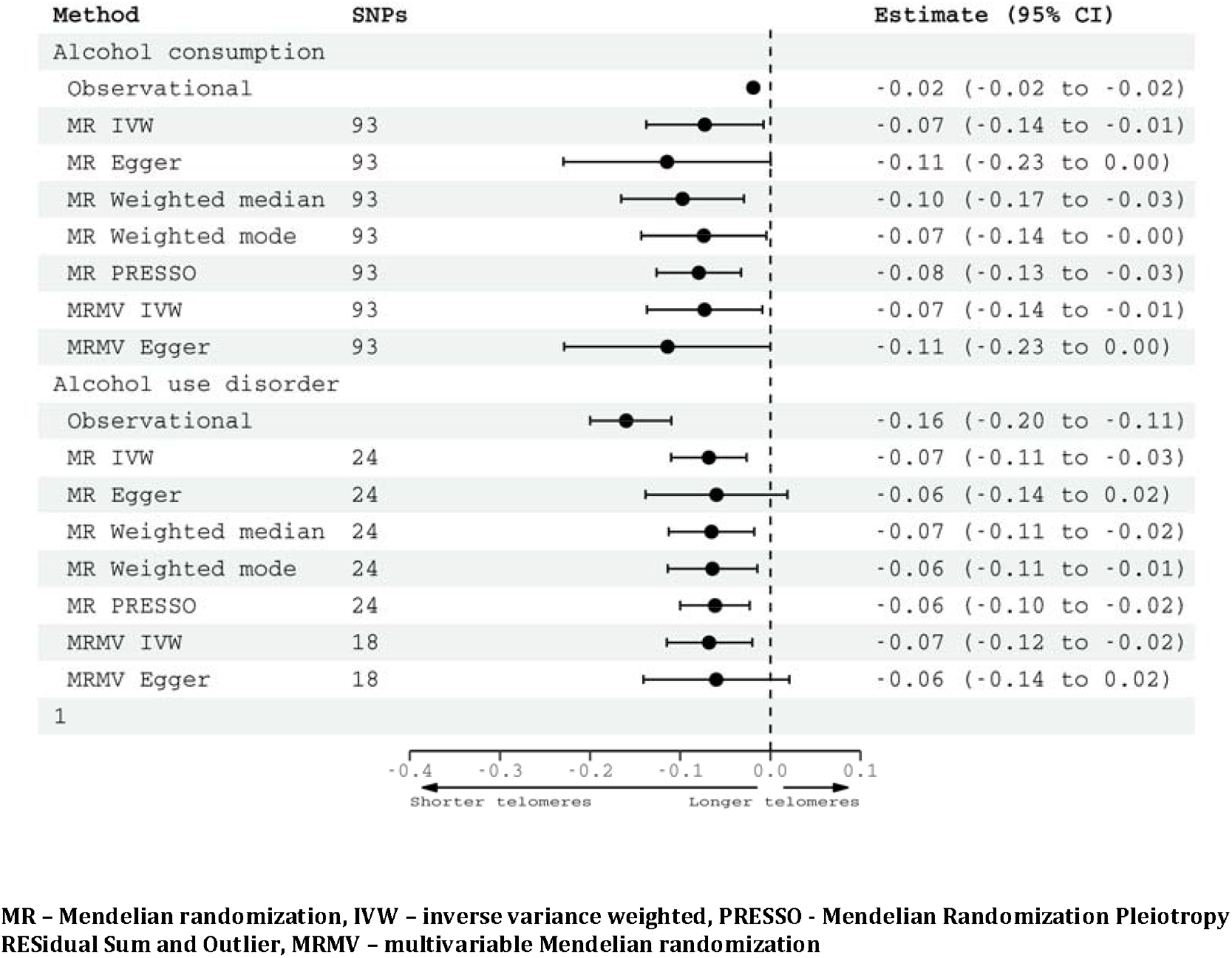
Multivariable-adjusted observational estimates (in 298,628 UKB participants) and Mendelian randomization estimates (two-sample design) for the association of genetically predicted alcohol consumption and alcohol use disorder with telomere length. Effect estimates for alcohol consumption are per SD increase in genetically-predicted log-transformed alcoholic drinks per week, and for AUD having a diagnosis of AUD.

In the AUD MR, using as instrumental variables 24 SNPs robustly and independently associated with AUD, there was a significant association with telomere length (IVW beta=-0.07, 95% CI: -0.11 to -0.03 p=0.001). Again, the estimates were consistent across all MR methods (Figure 2). Associations between genetically predicted alcohol intake and telomere length persisted in the multivariable IVW analysis (beta= -0.07, -0.14 to -0.01, p=0.03), controlling for smoking. Similarly, associations between genetically predicted AUD and telomere length remained significant in the IVW multivariable MR controlling for smoking (beta= -0.07, 95% CI: -0.12 to -0.02, p=0.005).

We found no evidence of reverse causation. MR Steiger tests for both alcohol consumption (p=9.0 × 10^−86^) and alcohol use disorder (p=2.9 × 10^−131^) indicated true causal effect directionality. Furthermore, neither associations between genetically predicted telomere length and alcohol consumption (IVW beta= 0.001, -0.02 to 0.04, p=0.6) nor AUD (IVW beta= -0.02, -0.06 to 0.02, p=0.3) were significant (supplementary Figure 10-13).

In non-linear MR analyses, associations between genetically predicted alcohol consumption and telomere length were only significant in the highest two quantiles of (IV-free) alcohol consumption, equating to >17 units weekly (17-28 units IVW beta= -0.17, 95% CI -0.30 to -0.04, p=0.013; >28 units IVW beta= -0.20, 95% CI: -0.40 to -0.004, p=0.046)(Figure 3). In participants drinking smaller amounts, there was no association. The trend in estimates was significant, as deemed by a meta-regression of the stratum estimates on median alcohol consumption in each stratum (p=0.002). These results provide a higher degree of confidence in a potentially causal effect in moderate to heavy drinkers than light drinkers.

**Figure 3:**
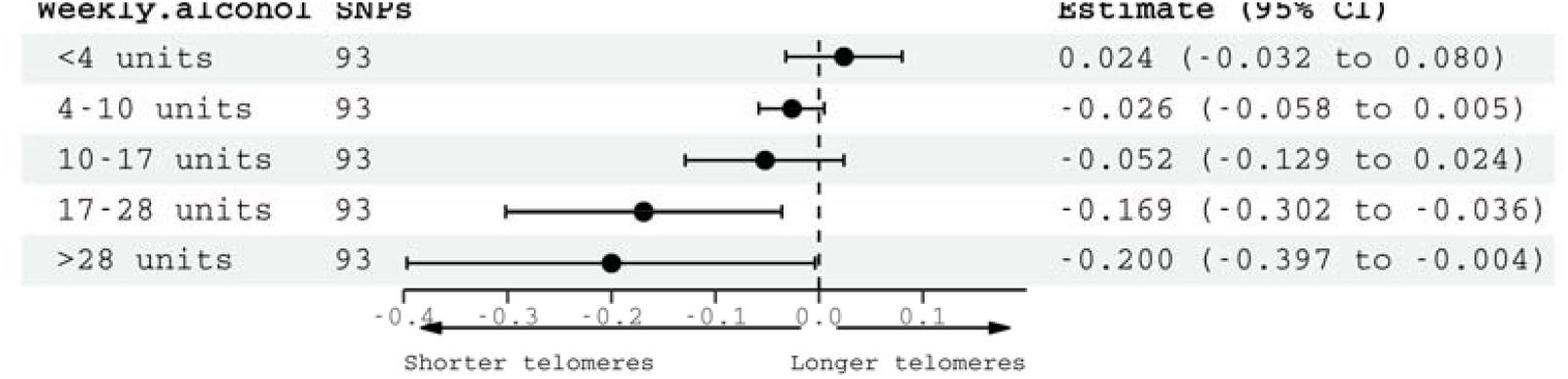
Non-linear Mendelian randomization showing associations between genetically predicted alcohol consumption and telomere length, stratified by weekly alcohol intake (IV-free exposure).

We found evidence for a causal effect of genetically predicted alcohol consumption on telomere length in current drinkers (IVW beta=-0.08, -0.15 to - 0.007, p=0.03) but not in never drinkers (beta=0.001, -0.02 to 0.02, p=0.9) (Figure 4). Similarly, a causal effect of AUD on telomere length was found in current drinkers (IVW beta=-0.10, -0.15 to -0.05, p<0.001) but not in never drinkers (IVW beta=0.18, -0.01 to 0.36, p=0.06) (Figure 5).

**Figure 4:**
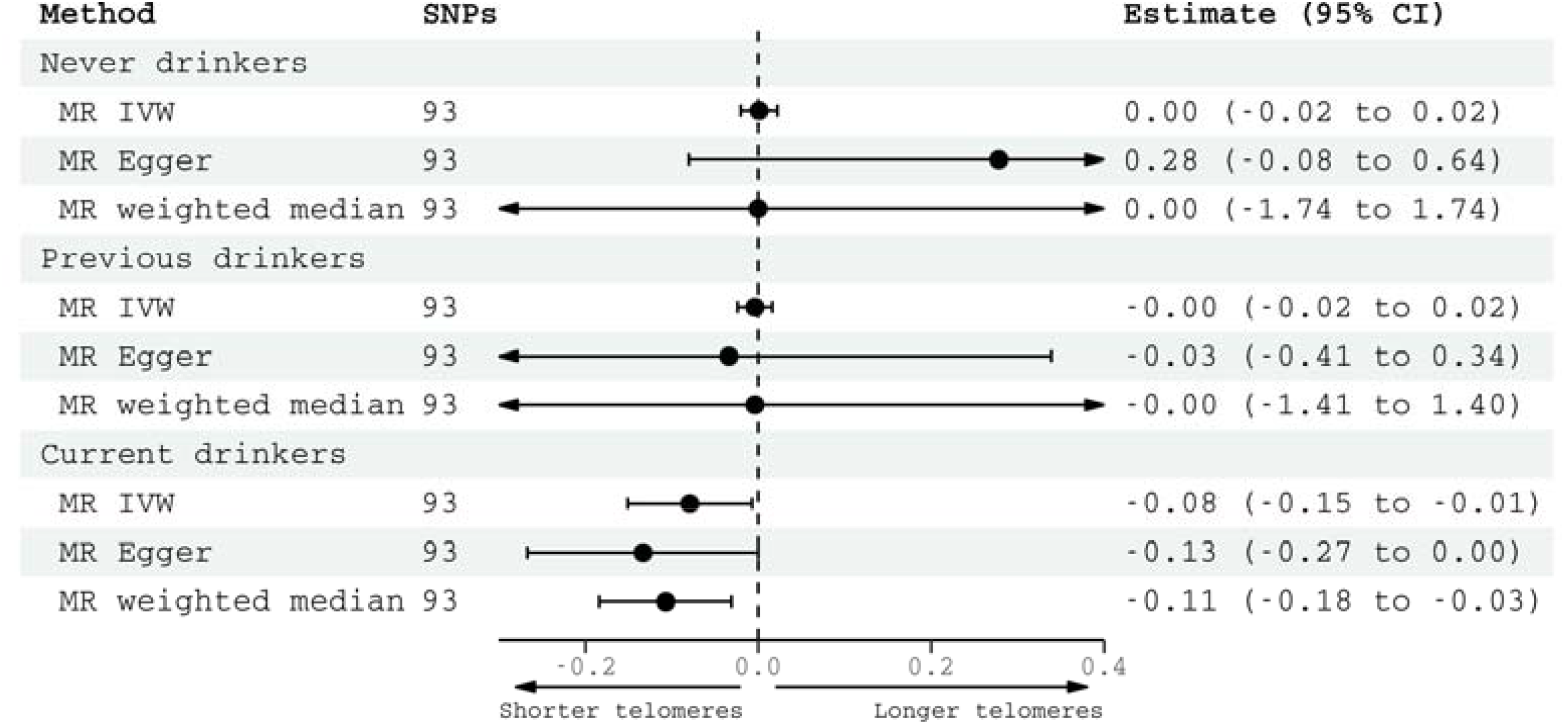
Negative controls for alcohol consumption. Causal estimates for alcohol consumption on telomere length generated by Mendelian randomization analyses according to alcohol status (n=12,433 never drinkers, n=13,582 previous drinkers, n=369,097 current drinkers. Effect estimates are per SD increase in genetically-predicted log-transformed alcoholic drinks per week.

**Figure 5:**
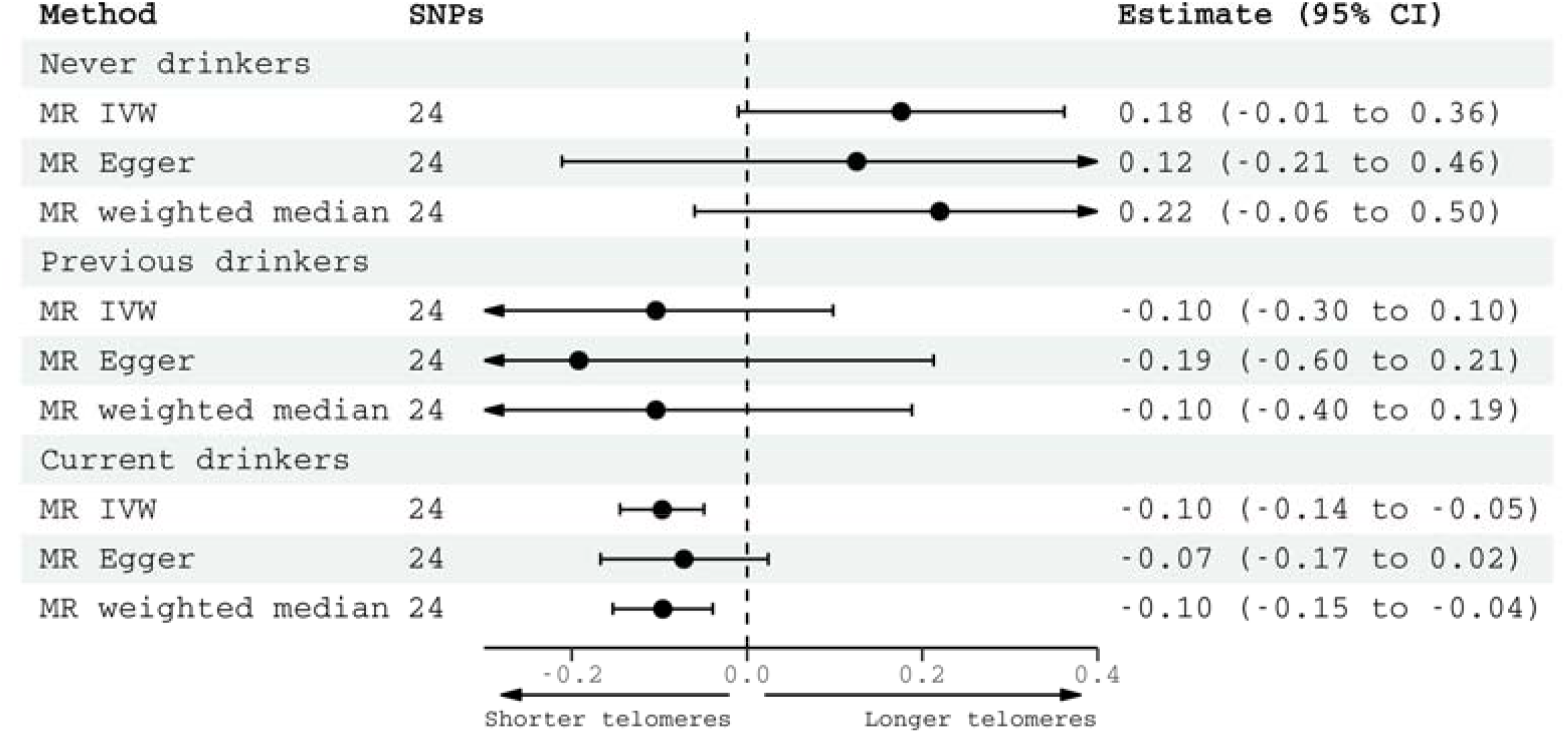
Negative controls for alcohol use disorder. Causal estimates for alcohol use disorder on telomere length generated by Mendelian randomization analyses according to alcohol status (n=12,433 never drinkers, n=13,582 previous drinkers, n=369,097 current drinkers). Effect estimates represent associations of a genetically predicted diagnosis of AUD vs. no diagnosis.

## Discussion

### Key findings

Using observational and MR approaches we observed consistent associations between two alcohol phenotypes, alcohol consumption and AUD, and shorter telomere length. Non-linear analyses were suggestive of a threshold relationship between alcohol intake and telomere length.

### Discussion of findings

Estimates for associations between genetically predicted alcohol consumption and telomere length were broadly consistent across the four MR methods employed. Whilst IV assumptions can never be tested empirically for each SNP, each method allows for different violations of the MR assumptions. Therefore consistent results across methods give greater confidence about the plausibility of the assumptions. The strongest association between genetically predicted alcohol consumption/AUD and telomere length was for rs1229984. The finding is biologically plausible, as this SNP is within an alcohol metabolism gene, *ADH1B*. It could result from greater power to detect a causal effect, as rs1229984 had the strongest associations with a broad range of alcohol use traits of any instrument used.

Again, for AUD all MR methods gave consistent causal estimates. Alcohol consumption and alcohol use disorder are distinct phenotypes, with only partial overlap in their genetic associations. The reasons for this are unclear [27]. But unlike the quantity-frequency measure AUDIT-C, AUD shows strong genetic correlation with a range of psychiatric disorders and negative medical outcomes [28]. AUD heritability could be partially explained by inherited personality traits, such as impulsivity [29] or sensation-seeking [30] which are less relevant for lower intakes. Overlap with genetic risk to psychiatric disorders such as depression [31] could also be a factor, or even propensity to physiological side effects following large quantities of alcohol.

To contextualize the effect size, in the observation analysis, drinking >29 units of alcohol weekly (compared to <6 units) was equivalent to 1-2 years of age-related change on telomere length. The MR effect sizes were greater - 1 S.D. higher genetically-predicted log-transformed alcoholic drinks weekly was equivalent to 3 years of aging. Possible explanations for greater associations in MR analyses are that these may capture the cumulative effects of lifetime propensity to drinking, and be subject to less confounding that observational estimates.

Significant associations between genetically predicted alcohol and telomere length were only found in current drinkers, providing support that the only path from the genetic variants to LTL is through alcohol. Furthermore, the strength of evidence for a causal effect of alcohol on telomere length was greater in heavier drinkers. This finding suggests a threshold effect, in that a necessary minimum amount of alcohol consumption is required to damage telomeres. Similar relationships with alcohol have been described for other health outcomes [32]. Additionally, multivariable MR analysis suggested that alcohol’s effects are direct and not mediated or confounded by smoking.

One mechanism by which alcohol could exert an influence on telomere length could be via oxidative stress and inflammation. Oxidative stress has been demonstrated in vitro and in vivo to affect telomere length [33]. Ethanol metabolism can produce reactive oxygen species and reactive nitrogen species [34, 35]. In addition, ethanol can reduce levels of critical cellular antioxidants, such as glutathione [36, 37], compounding the oxidative stress.

### Strengths

Strengths of this study include the triangulation of observational and MR approaches. The observational analysis is the largest to date. No previous MR analysis has been undertaken. Two alcohol traits, alcohol consumption and AUD were examined. Univariate and multivariate two-sample MR analyses were performed, as well as individual-level data interrogated in a non-linear MR analysis. Genetic associations were extracted from the largest GWAS available for both exposures and outcomes published. Multiple sensitivity analyses, including use of negative controls and multivariate MR were undertaken to explore the robustness of the findings.

### Limitations

Some limitations need to be acknowledged. Genetic variants explained a low variance of alcohol traits. Despite this, our analysis had 85% statistical power to detect a 0.08 standard deviation change in telomere length for a 1 standard deviation increase in alcohol consumption. Due to size differences between groups, we had greater power to detect genetic associations in current drinkers compared to never drinkers. The observational analysis, use of negative controls, and non-linear MR analysis used self-report to determine alcohol intake. Although this is the only feasible method at scale, it may be subject to misclassification bias. MR estimates the causal effect of lifetime exposure to alcohol. Hence estimates do not necessarily equate to effect sizes if alcohol intake were modified following an intervention during adult life. Genetic associations with alcohol and telomere length were calculated in those with European ancestry and therefore may not apply to other populations with different ancestry groups. Two sample MR assumes that the two populations are broadly similar. UK Biobank is likely subject to a healthy volunteer bias. Prevalence of alcohol dependence in UKB was much lower than general population estimates [37]. This likely reflects cases being defined on the basis of ICD codes in linked health records, which would capture only the most severe cases. MR techniques rely on a number of assumptions which we have tried to test where possible, but residual uncertainty inevitably remains. Finally, telomere length was measured in leucocytes, but the extent to which this reflects other organ tissues is not clear [38].

### Conclusions

In conclusion, associations between alcohol traits, and genetically predicted alcohol traits, and telomere length were found. Non-linear analyses suggested that a threshold alcohol intake may be necessary to impact telomere length. These findings lend support to alcohol being a causal determinant of telomere length. Multiple sensitivity analyses to assess assumptions of the estimation methods offer a degree of confidence to their plausibility. Such findings provide further evidence of the harmful effects of alcohol on biological aging. Furthermore, they may inform strategies to prevent alcohol- and aging-related diseases, including neurodegenerative conditions.

## Data Availability

Data is available on successful application to the UK Biobank.

## Notes

**<u>Funding:</u>** AT is supported by a Wellcome Trust fellowship (216462/Z/19/Z). KPE is funded by the UK Medical Research Council (G1001354 & MR/K013351/1) and the European Commission (Horizon 2020 732592). SB is supported by a Sir Henry Dale Fellowship jointly funded by the Wellcome Trust and the Royal Society (204623/Z/16/Z). This work was also supported by the Li Ka Shing Centre for Health Information and Discovery, NIH grant (TN: R01EB026859), the National Institute for Health Research Cambridge Biomedical Research Centre (BRC-1215-20014), and a Wellcome Trust award (TN: 100309/Z/12/Z).

### Competing Interest Statement

The authors have declared no competing interest.

### Funding Statement

AT is supported by a Wellcome Trust fellowship (216462/Z/19/Z).
KPE is funded by the UK Medical Research Council (G1001354 & MR/K013351/1) and the European Commission (Horizon 2020 732592). SB is supported by a Sir Henry Dale Fellowship jointly funded by the Wellcome Trust and the Royal Society (204623/Z/16/Z). This work was also supported by the Li Ka Shing Centre for Health Information and Discovery, NIH grant (TN: R01EB026859), the National Institute for Health Research Cambridge Biomedical Research Centre (BRC-1215-20014), and a Wellcome Trust award (TN: 100309/Z/12/Z).

### Author Declarations

UKB approval was obtained from the North West Multi-Centre Research Ethics Committee and the Patient Information Advisory Group. The UKB Board and Access Sub-Committee and the Biobank Ethics and Governance Council reviewed our data access application.

