## Supplementary methods and tables for "Alcohol consumption and telomere length: observational and Mendelian randomization approaches"

Alcohol use disorder (AUD) cases were defined by the presence of a relevant ICD-9 (303, 30301, 30302, 30303, 3039, 30391, 30392, 30393) or ICD-10 (F102, F1021, F1022, F1023, F1024, F1025, F1026, F1027, F1028, F1029).

Non-linear MR: A meta-regression was performed of the linear MR estimates in each stratum against mean alcohol consumption in that stratum. Then the fractional polynomial method was used to assess whether a non-linear model fitted this meta-regression better than a linear model [27].

**Supplementary results**

|  | **Estimate (95% CI)** |
| --- | --- |
| **Q2 vs. Q1 drinker** | 0.01 (-0.003, 0.02) |
| **Q3 vs. Q1 drinker** | 0.01^**^ (0.0004, 0.02) |
| **Q4 vs. Q1 drinker** | -0.01 (-0.02, 0.01) |
| **Q5 vs. Q1 drinker** | -0.05^***^ (-0.07, -0.04) |
| **Never vs. Q1 drinker** | 0.02 (-0.001, 0.04) |
| **Previous vs. Q1 drinker** | -0.03^*^ (-0.05, 0.001) |
| **Age, *years*** | -0.02^***^ (-0.02, -0.02) |
| **Sex** | -0.16^***^ (-0.17, -0.15) |
| **Ethnicity^1^:**  ***Mixed*** | -0.05 (-0.47, 0.38) |
| ***Asian*** | -0.16 (-0.61, 0.28) |
| ***Black*** | 0.28 (-0.36, 0.93) |
| ***Chinese*** | 0.37^***^ (0.23, 0.51) |
| ***Other*** | 0.17^**^ (0.04, 0.29) |
| ***British*** | 0.01 (-0.11, 0.14) |
| ***Irish*** | 0.08 (-0.04, 0.20) |
| ***Other white*** | 0.13^**^ (0.01, 0.25) |
| ***White and Black Carribean*** | 0.11 (-0.05, 0.27) |
| ***White and Black African*** | 0.14 (-0.04, 0.31) |
| ***White and Asian*** | 0.10 (-0.05, 0.25) |
| ***Other mixed*** | 0.13^*^ (-0.01, 0.28) |
| ***Indian*** | -0.01 (-0.13, 0.12) |
| ***Pakistani*** | 0.02 (-0.12, 0.15) |
| ***Bangladeshi*** | -0.11 (-0.32, 0.10) |
| ***Other Asian*** | 0.06 (-0.08, 0.19) |
| ***Carribean*** | 0.42^***^ (0.30, 0.55) |
| ***African*** | 0.42^***^ (0.29, 0.55) |
| ***Other Black*** | 0.28^**^ (0.01, 0.55) |
| **Constant** | 1.43^***^ (1.31, 1.55) |
| **N** | 299,980 |
| **R^2^** | 0.05 |
| **Adjusted R^2^** | 0.05 |
| **Residual Std. Error** | 0.97 (df = 299952) |
| **F Statistic** | 619^***^ (df = 27; 299952) |
| ^*^p < .1; ^**^p < .05; ^***^p < .01  ^1^ Reference group White | |

**Alcohol quartiles:**

Quintile 1 (Q1) <5.79 units weekly

Quintile 2 (Q2) 5.79-10.8 units weekly

Quintile 3 (Q3) 10.8-17.2 units weekly

Quintile 4 (Q4) 17.2-28.9 units weekly

Quintile 5 (Q5) >28.9 units weekly

**Table 1: Regression model examining for association between alcohol consumption (categorical) and telomere length.**

|  | **Estimate (95% CI)** |
| --- | --- |
| **Alcohol dependence** | -0.18^***^ (-0.19, -0.14) |
| **Age, years** | -0.02^***^ (-0.02, -0.02) |
| **Sex** | -0.17^***^ (-0.18, -0.16) |
| **Ethnicity^1^:**  ***Mixed*** | -0.04 (-0.46, 0.38) |
| ***Asian*** | -0.15 (-0.59, 0.29) |
| ***Black*** | 0.28 (-0.36, 0.93) |
| ***Chinese*** | 0.37^***^ (0.24, 0.51) |
| ***Other*** | 0.17^***^ (0.04, 0.29) |
| ***British*** | 0.01 (-0.11, 0.13) |
| ***Irish*** | 0.07 (-0.05, 0.19) |
| ***Other white*** | 0.13^**^ (0.01, 0.25) |
| ***White and Black Carribean*** | 0.11 (-0.05, 0.26) |
| ***White and Black African*** | 0.13 (-0.04, 0.31) |
| ***White and Asian*** | 0.10 (-0.05, 0.24) |
| ***Other mixed*** | 0.13^*^ (-0.01, 0.27) |
| ***Indian*** | -0.01 (-0.13, 0.12) |
| ***Pakistani*** | 0.03 (-0.11, 0.16) |
| ***Bangladeshi*** | -0.10 (-0.31, 0.11) |
| ***Other Asian*** | 0.06 (-0.07, 0.19) |
| ***Carribean*** | 0.42^***^ (0.30, 0.55) |
| ***African*** | 0.42^***^ (0.29, 0.55) |
| ***Other Black*** | 0.28^**^ (0.01, 0.55) |
| **Constant** | 1.43^***^ (1.31, 1.56) |
| **N** | 299,980 |
| **R^2^** | 0.05 |
| **Adjusted R^2^** | 0.05 |
| **Residual Std. Error** | 0.97 (df = 299957) |
| **F Statistic** | 755^***^ (df = 22; 299957) |
| ^*^p < .1; ^**^p < .05; ^***^p < .01 | |

^1^ Reference group White

**Table 2: Regression model examining for association between alcohol use disorder (defined using ICD codes in linked HES records) and telomere length.**

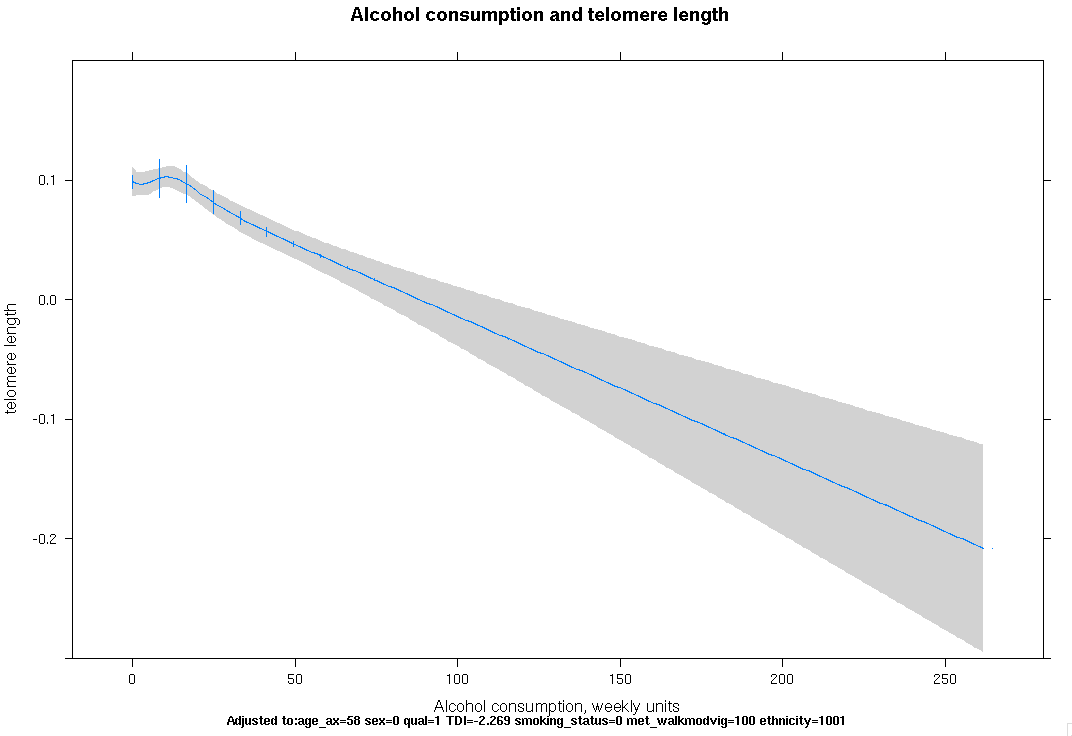

**Figure 1: Observational association of alcohol consumption with telomere length in n=298,628 UK Biobank participants. Restricted cubic splines fitted to alcohol with 5 knots. Regression models adjusted for: age, sex, ethnicity.**

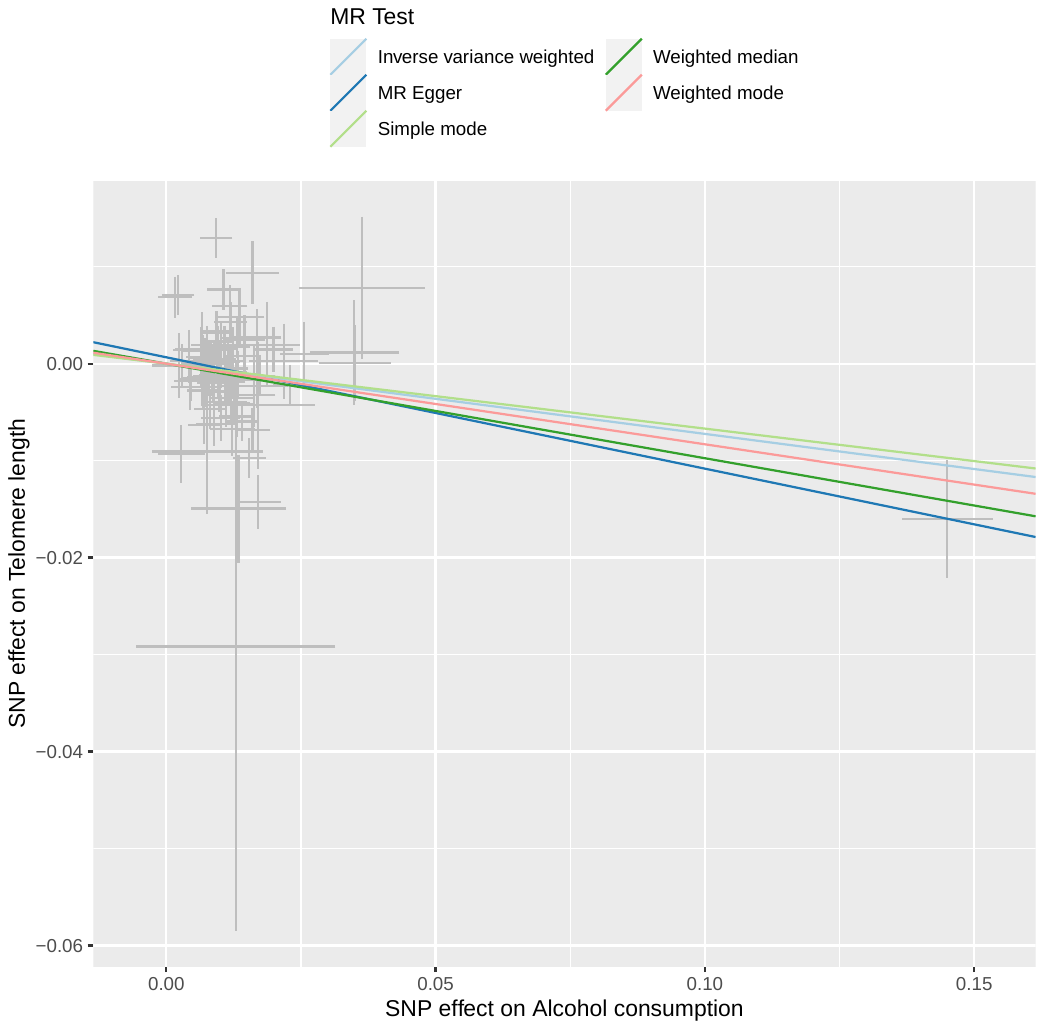

IVW beta= -0.07, p=0.0328

MR-Egger beta= -0.11, p=0.054

Weighted median beta= -0.10, p=0.005

Weighted mode beta= -0.08, p=0.023

**Figure 2: Associations between genetically predicted alcohol consumption and telomere length from two-sample Mendelian randomization. Graph shows the strength of association between telomere length and alcohol consumption SNPs on the y-axis against the alcohol consumption associations from previous genome-wide association studies for each SNP on the x-axis. A non-zero gradient to the lines indicates evidence for causality of alcohol on telomere length.**

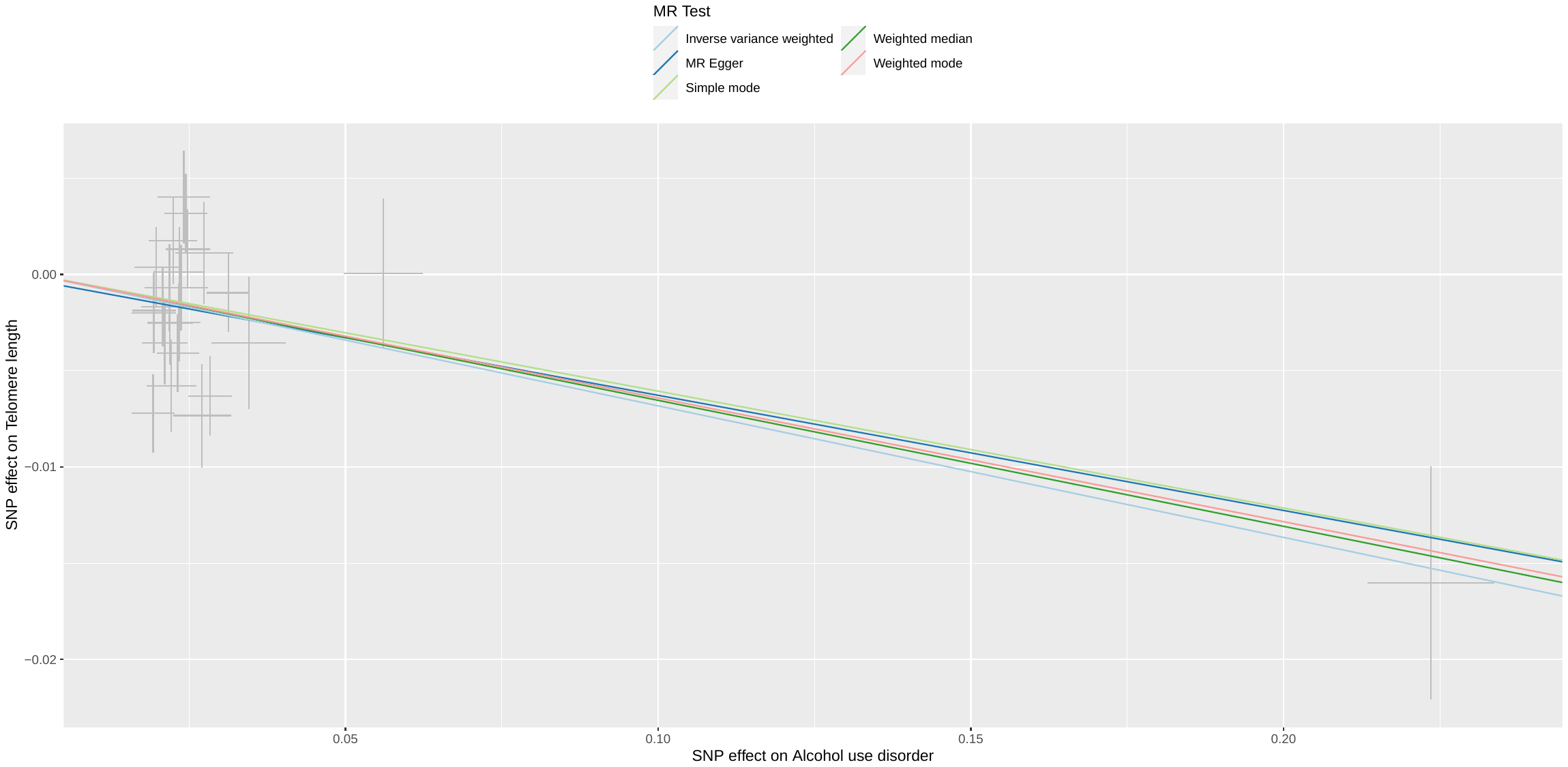

IVW beta= -0.07, p=0.001

MR-Egger beta= -0.06, p=0.15

Weighted median beta= -0.07, p= 0.007

Weighted mode beta= -0.06, p=0.02

**Figure 3: Associations between genetic propensity to alcohol use disorder (AUD) and telomere length generated from two sample Mendelian randomization. Graph shows the strength of association between telomere length and AUD SNPs on the y-axis against the AUD associations from previous genome-wide association studies for each SNP on the x-axis. A non-zero gradient to the lines indicates evidence for causality of AUD on telomere length.**

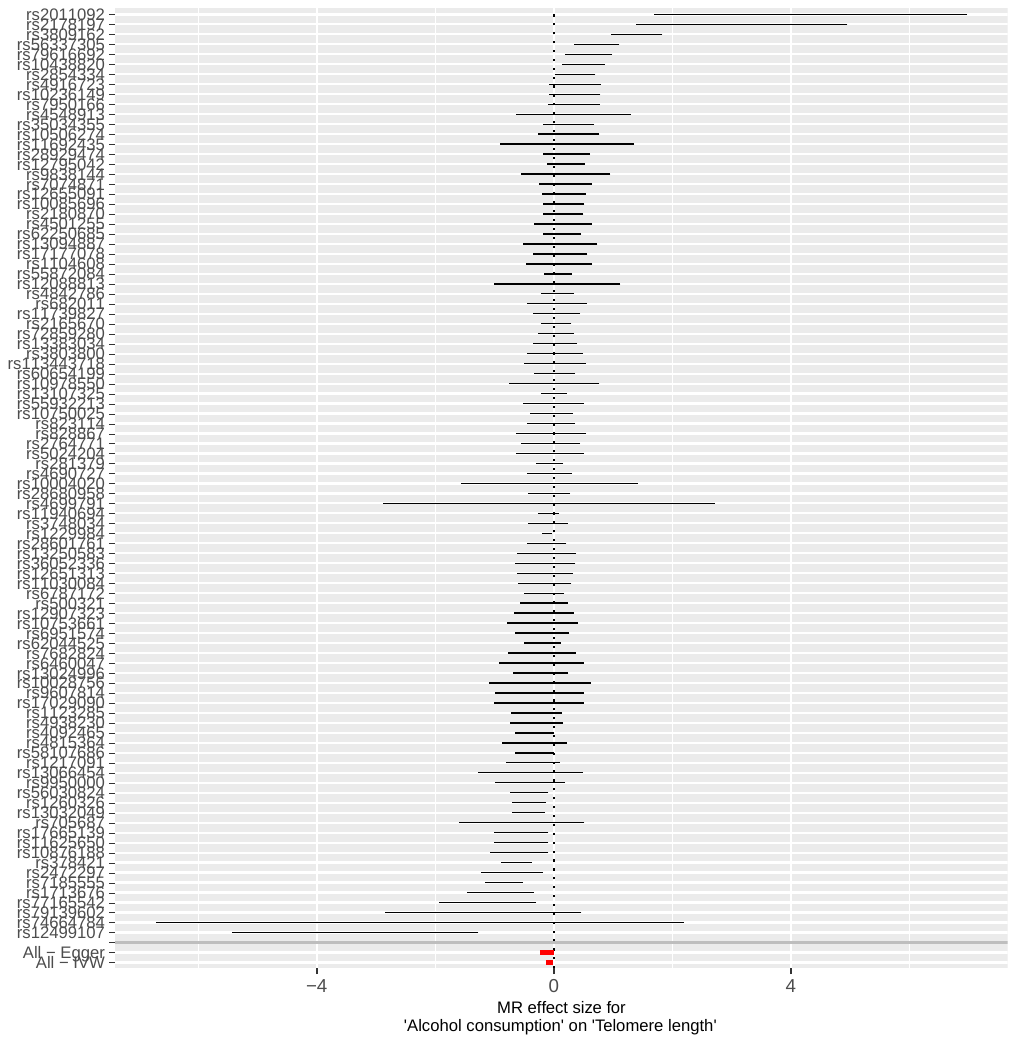

**Figure 4: Forest plot showing Mendelian randomization estimates for association of genetically predicted alcohol consumption and telomere length for each SNP.**

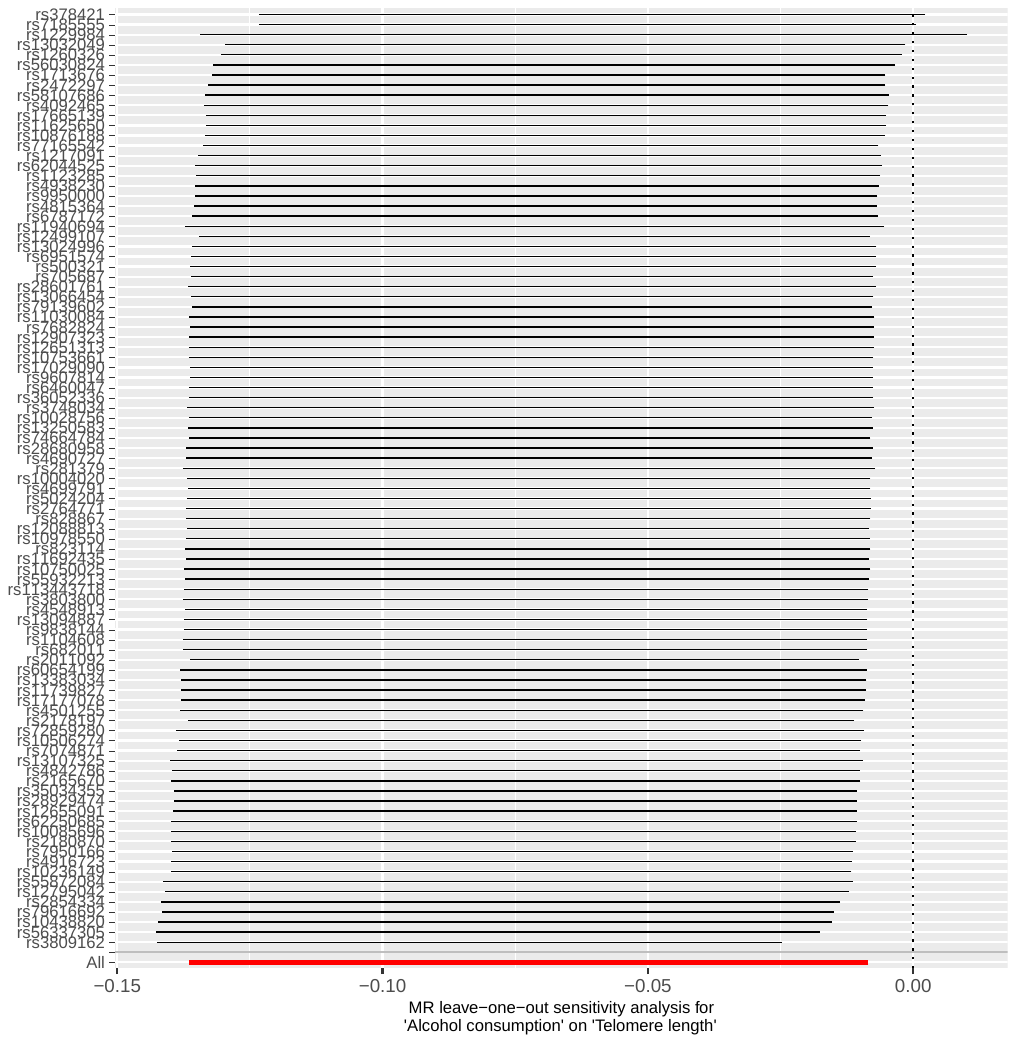

**Figure 5: Leave one out plot for genetically predicted alcohol consumption association with telomere length.**

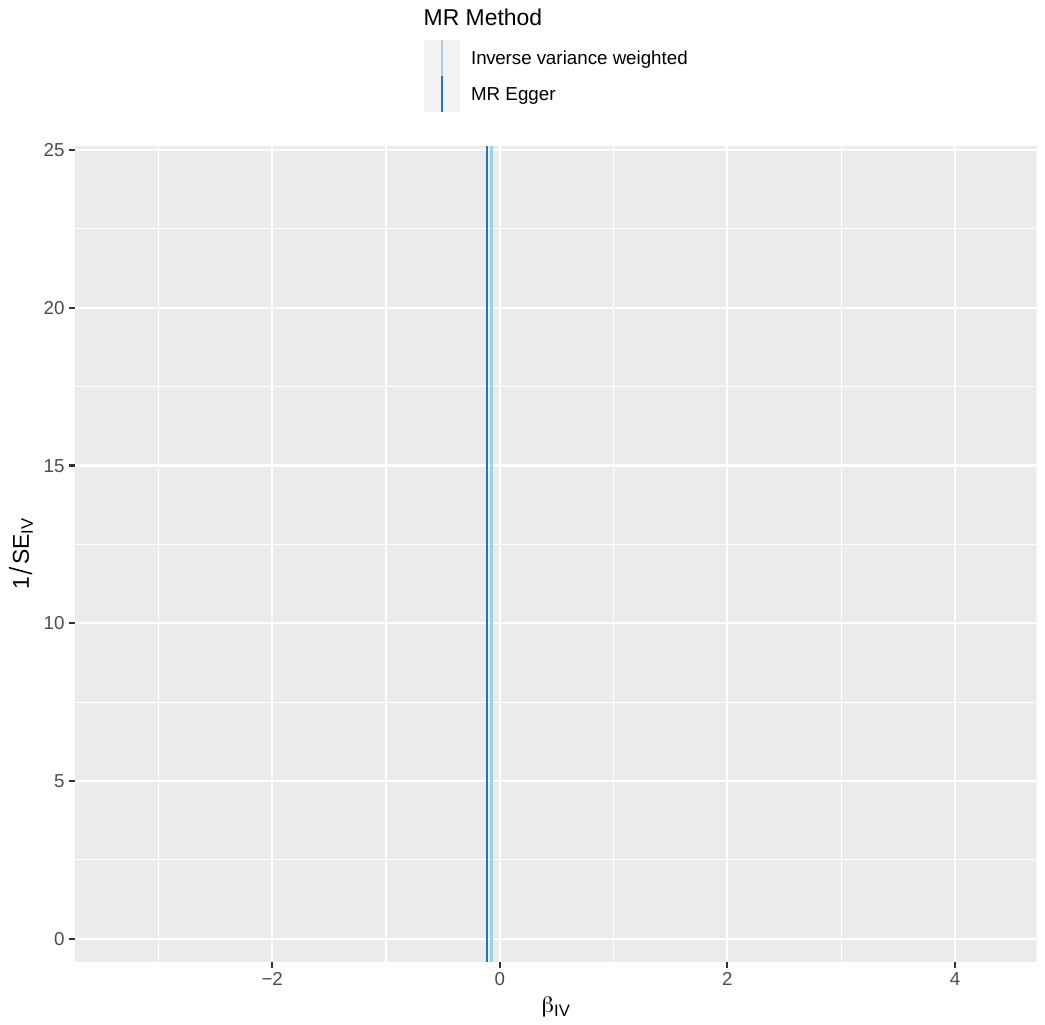

**Figure 6: Funnel plot of genetically predicted alcohol consumption association with telomere length.**

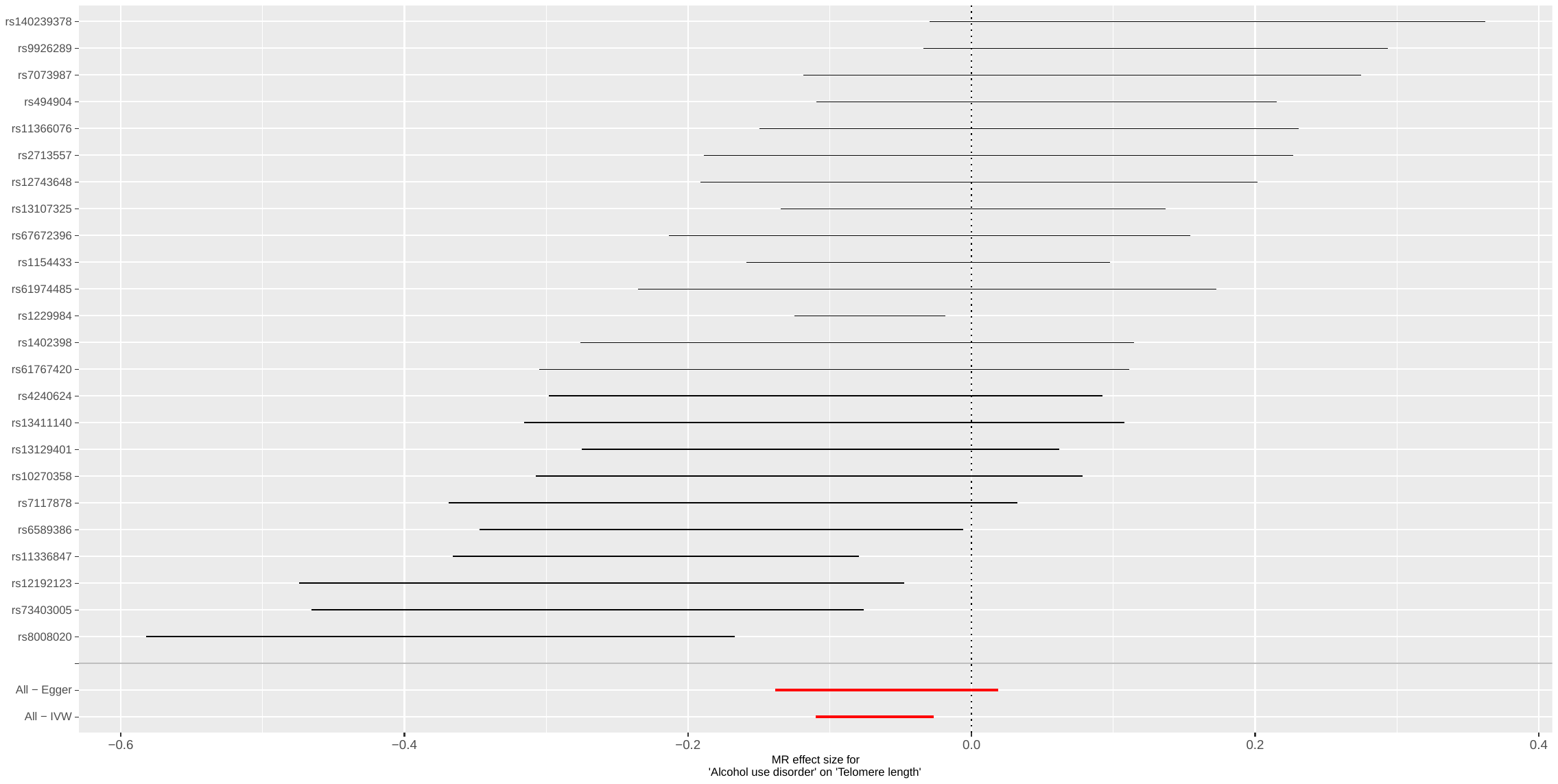

**Figure 7: Forest plot showing Mendelian randomization estimates for genetically predicted alcohol use disorder association with telomere length for each SNP.**

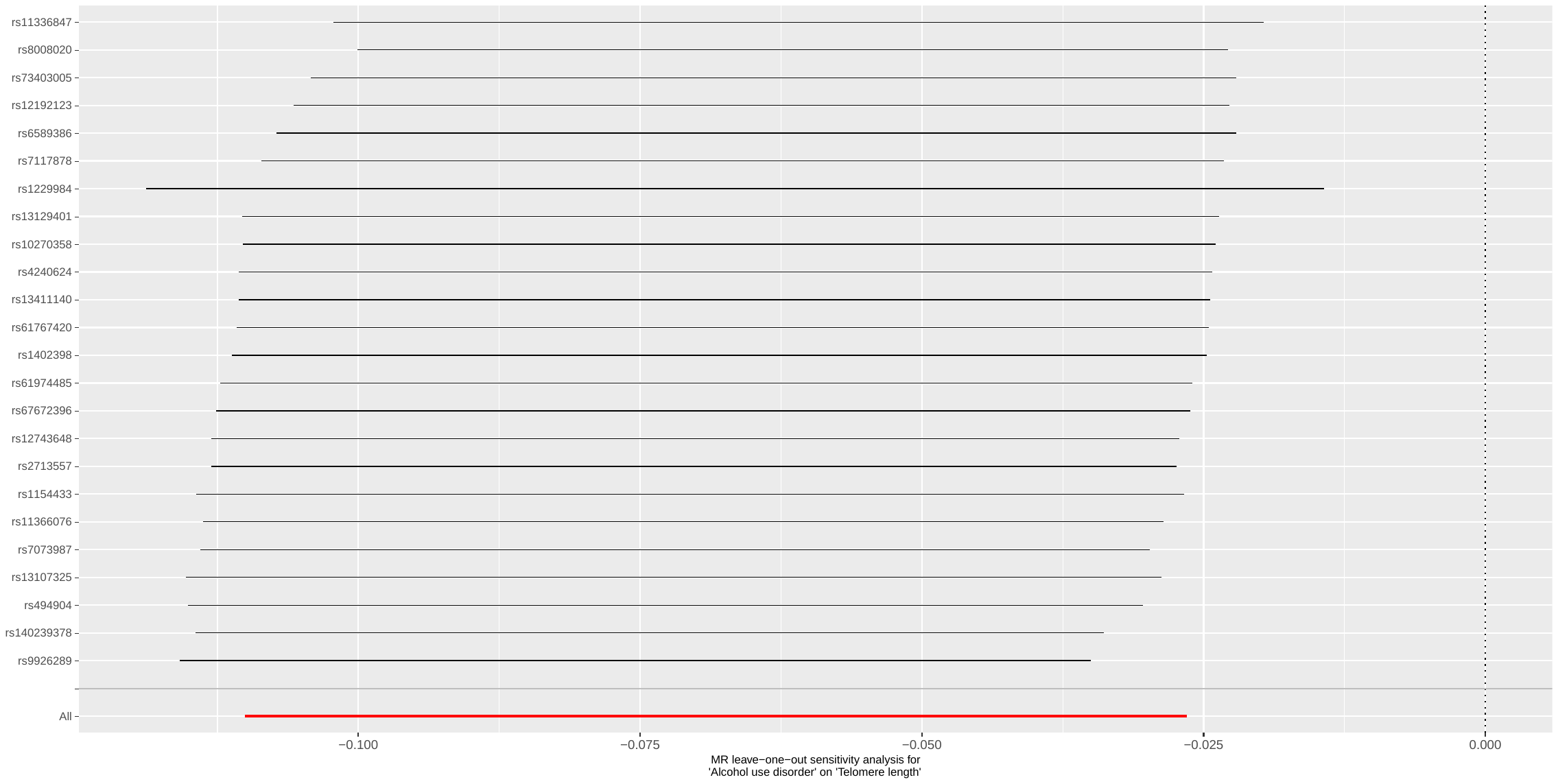

**Figure 8: Leave one out plot for genetically predicted alcohol use disorder association with telomere length**

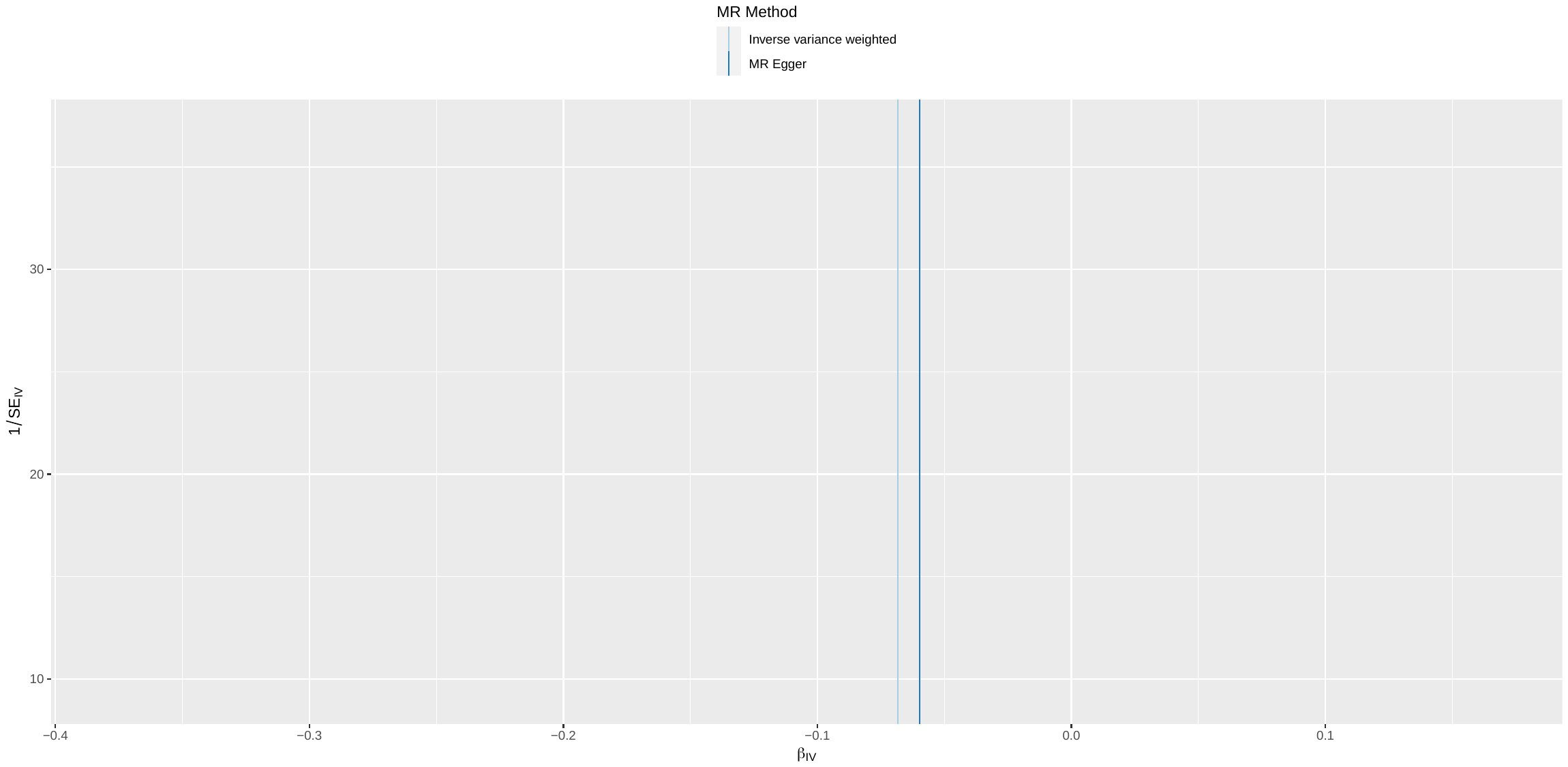

**Figure 9: Funnel plot for association of genetically predicted alcohol use disorder and telomere length.**

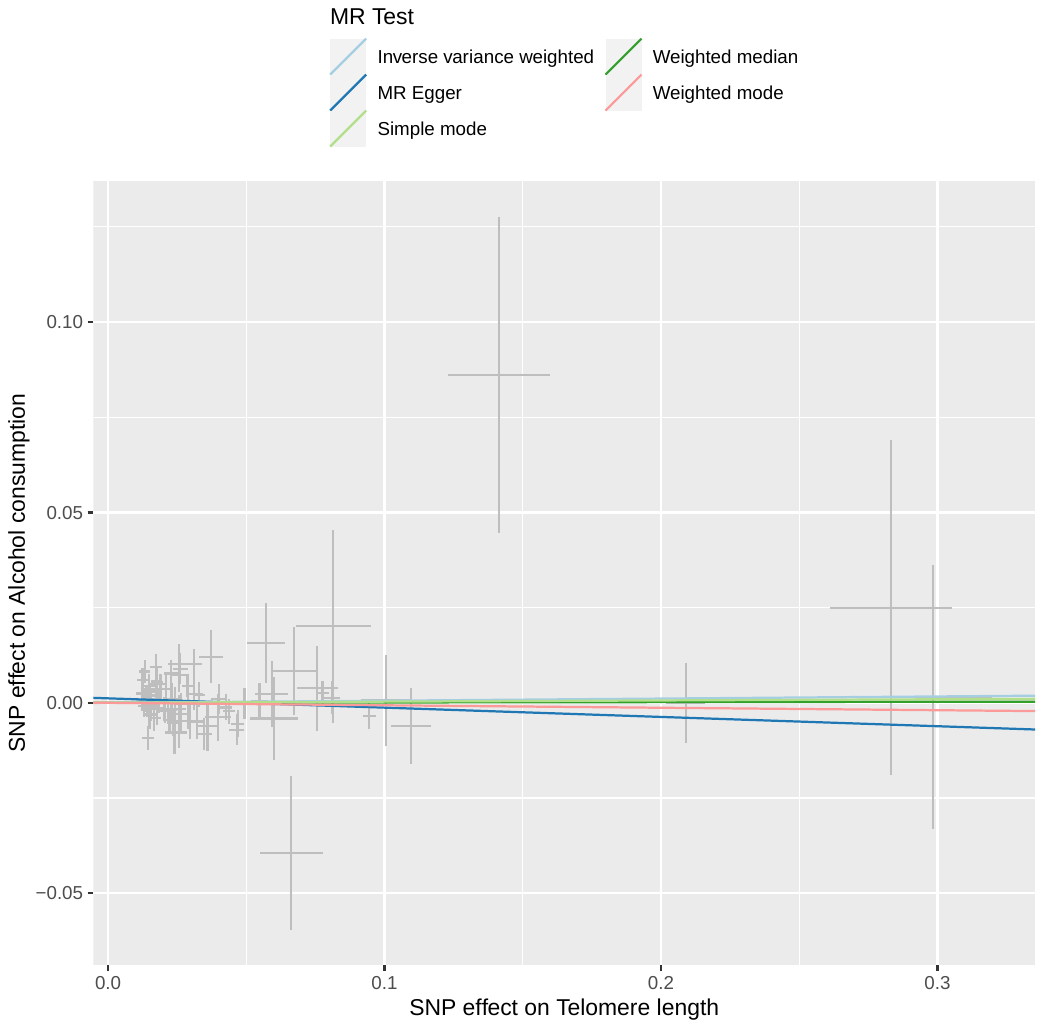

IVW beta= 0.006, p=0.7

MR-Egger beta=-0.02, p=0.3

Weighted mode beta=-0.006, p=0.7

**Figure 10: Two sample Mendelian randomization showing association of genetically predicted telomere length and alcohol consumption (checking for reverse causation). Graph shows the strength of association between alcohol consumption and telomere SNPs on the y-axis against the telomere associations from previous genome-wide association studies for each SNP on the x-axis. A non-zero gradient to the lines indicates evidence for causality of telomere length on alcohol consumption. 85 SNPs included as instruments with complete outcome associations (n=5 SNPs palindromic with intermediate allele frequencies excluded).**

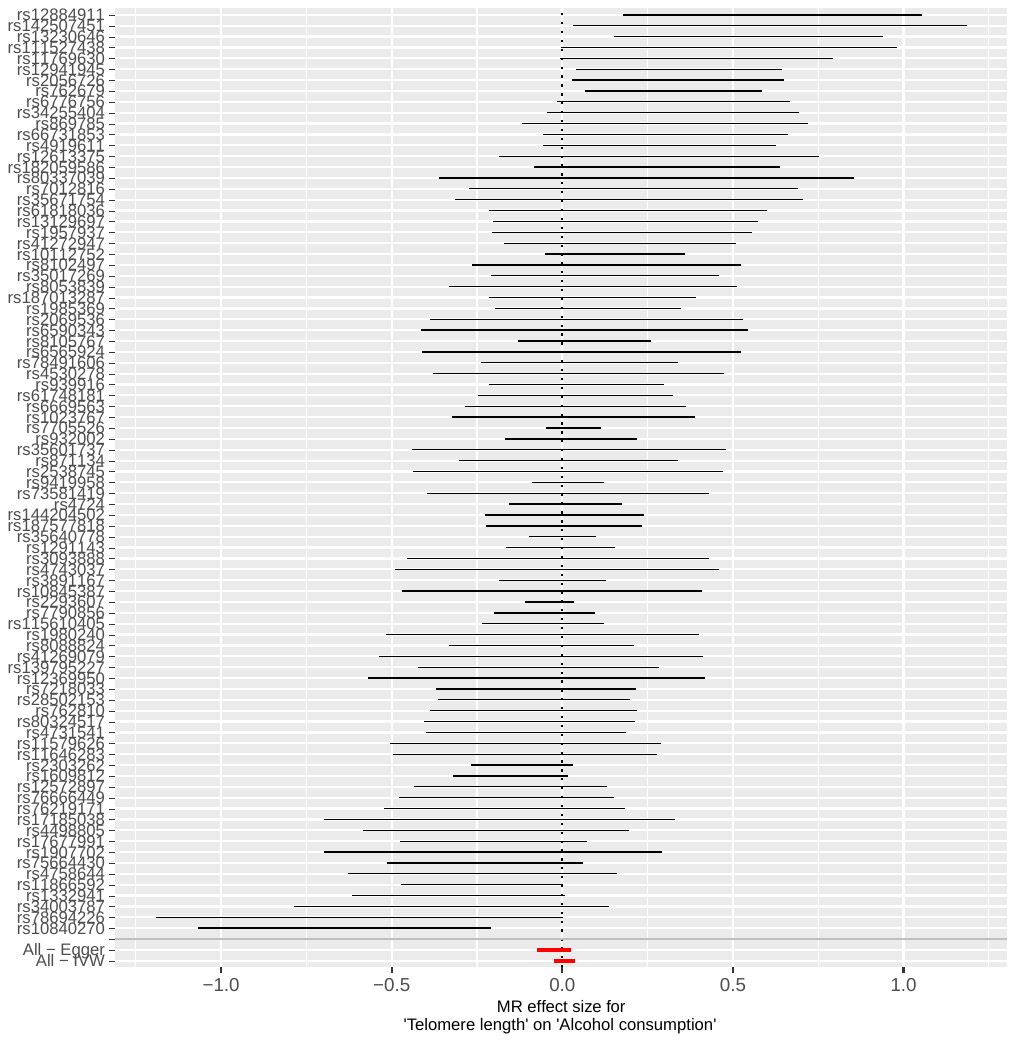

**Figure 11: Forest plot showing estimates for genetically predicted telomere length and alcohol consumption associations for each SNP.**

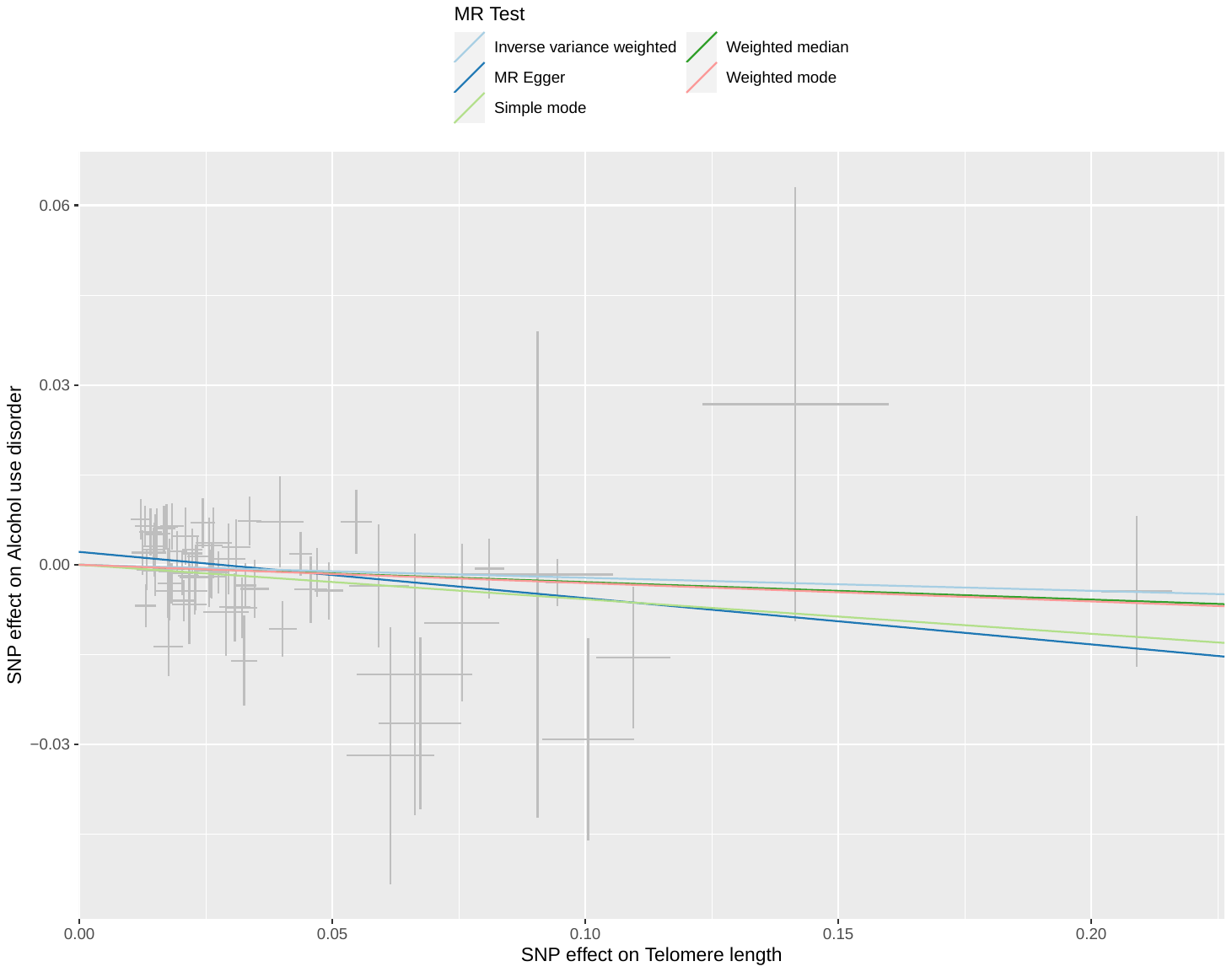

**Figure 12: Scatterplot showing genetically predicted telomere length and alcohol use disorder associations. N=67 SNPs with available outcome data (n=1 palindromic SNPs with intermediate allele frequencies excluded).**

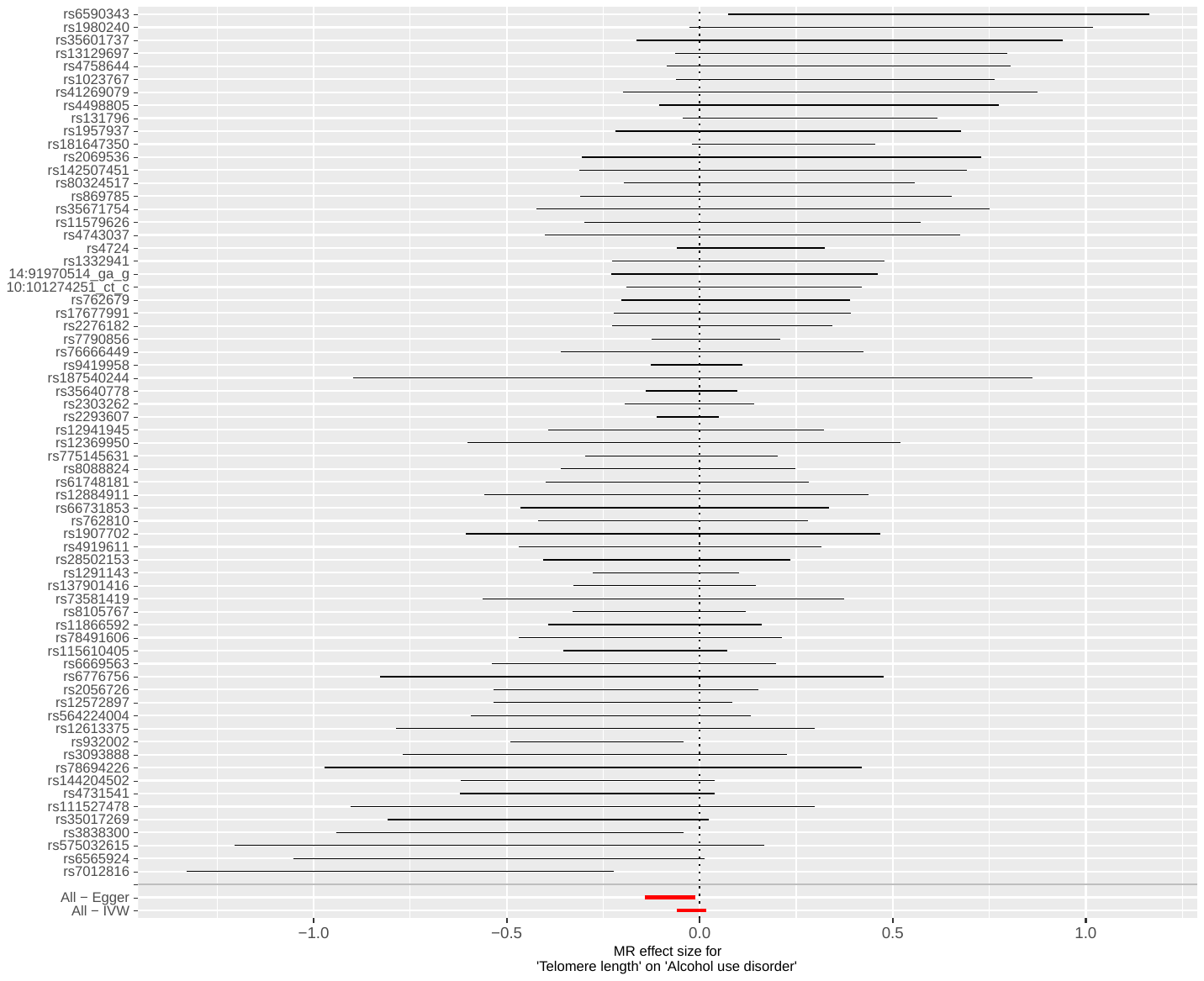

**Figure 13: Forest plot showing SNP estimates for associations between genetically predicted telomere length and alcohol use disorder.**

| **Non-linearity test** | **P value** |
| --- | --- |
| Fractional polynomial degree | 0.9 |
| Fractional polynomial non-linearity | 0.1 |
| Quadratic | 0.1 |
| Cochran Q | 0.5 |

**Table 3: Non-linearity tests for non-linear Mendelian randomization. The best-fitting polynomial of degree 1 for the relationship between alcohol and telomere length had power 2.**
